# A Consensus Definition of Misophonia: Using a Delphi Process to Reach Expert Agreement

**DOI:** 10.1101/2021.04.05.21254951

**Authors:** Susan Swedo, David M. Baguley, Damiaan Denys, Laura J. Dixon, Mercede Erfanian, Alessandra Fioretti, Pawel J. Jastreboff, Sukhbinder Kumar, M. Zachary Rosenthal, Romke Rouw, Daniela Schiller, Julia Simner, Eric A. Storch, Steven Taylor, Kathy R. Vander Werff, Sylvina M. Raver

## Abstract

Misophonia is a disorder of decreased tolerance to specific sounds or their associated stimuli that has been characterized using different language and methodologies. The absence of a common understanding or foundational definition of misophonia hinders progress in research to understand the disorder and develop effective treatments for individuals suffering from misophonia. From June 2020 through January 2021, a project was conducted to determine whether a committee of experts with diverse expertise related to misophonia could develop a consensus definition of misophonia. An expert committee used a modified Delphi method to evaluate candidate definitional statements that were identified through a systematic review of the published literature. Over four rounds of iterative voting, revision, and exclusion, the committee made decisions to include, exclude, or revise these statements in the definition based on the currently available scientific and clinical evidence. A definitional statement was included in the final definition only after reaching consensus at 80% or more of the committee agreeing with its premise and phrasing. The results of this rigorous consensus-building process were compiled into a final definition of misophonia that is presented here. This definition will serve as an important step to bring cohesion to the growing field of researchers and clinicians who seek to better understand and support individuals experiencing misophonia.

## Introduction

Misophonia was named and described in the early 2000’s (Jastreboff and Jastreboff, 2001; Jastreboff and Jastreboff, 2002) and has since gained scientific recognition and clinical identification across a wide variety of disciplines (e.g., audiology, neuroscience, occupational therapy, psychiatry, psychology). To the layperson, misophonia could be narrowly understood as a strong dislike of certain sounds, such as chewing. However, despite a common appreciation that misophonia is present in individuals when specific sensory input, such as a particular sound, leads to strong emotional and physical responses, researchers and clinicians have characterized the disorder differently (e.g. Jastreboff and Jastreboff, 2002; Edelstein et al., 2013; Schröder et al., 2013; Wu et al., 2014; Brout et al., 2018). Scientific research investigating misophonia has been conducted for fewer than 20 years and the literature on misophonia has not yet surpassed 100 peer-reviewed papers. During this early phase of research, misophonia has been defined by different criteria with variable methods used to diagnose and assess symptom severity. As a result of this fundamental lack of consensus regarding how misophonia is defined and evaluated, comparisons between study cohorts are not possible, measurement tools have not been well psychometrically validated, and the field cannot rigorously assess the efficacy of different treatment approaches.

### Need for Consensus Definition

Beginning in June 2020, the Misophonia Research Fund (MRF) with guidance from the Milken Institute’s Center for Strategic Philanthropy initiated a project with the overall objective of identifying and publishing a consensus definition of misophonia for the scientific community. The MRF, an initiative of The REAM Foundation operated in partnership with the Center for Strategic Philanthropy, provides funding for medical research grants that seek to better understand misophonia, diagnose people who have the condition, and assess treatment strategies. A Scientific Advisory Board guides the MRF and identified the need to build a fundamental understanding of misophonia as an early strategic priority of the Fund. Any resulting definition from this consensus project is intended to be inclusive of current definitions of misophonia so that the consensus definition could capture the majority of individuals with misophonia. A standardized definition, adopted by clinicians and researchers, and understood by individuals with lived experience, is critical to create well-defined, streamlined cohorts for further study. It can serve as the foundation of future diagnostic criteria and validated diagnostic tools, and bring cohesion to the diverse and interdisciplinary misophonia research and clinical communities.

### About the Delphi Method

We sought to use an established and structured consensus-building process to develop a foundational definition. The Delphi method works on the assumption that group judgements are more valid than individual ones. The approach is an effective iterative process with repeated rounds of evidence evaluation and voting to determine consensus among a group of experts with different knowledge and varying levels of expertise about a particular topic (Gustafson et al., 1973; Murphy et al., 1998). Initially developed by researchers at the RAND Corporation (Dalkey, 1969), the Delphi method has been used in a variety of fields since the 1960’s to reach consensus. Variants of the original technique have been reliably used in medical science, healthcare, and mental health research for the purpose of defining foundational concepts, designing domains or criteria, and determining consensus definitions (Jorm, 2015; San et al., 2015; Eubank et al., 2016; Stern et al., 2018; Venkatesan et al., 2019).

Here, we employed a four-step Delphi method (Figure 1) that included two rounds of independent voting and asynchronous commentary through online surveys followed by a third round of expert discussion and voting via a virtual meeting. A fourth and final round of voting via online survey was held to finalize the details of the definition prose. While the original Delphi method did not include an interactive discussion among experts (Dalkey, 1969), we used a modified Delphi approach that included a voting round that consisted of a meeting for expert interaction (Gustafson et al., 1973). This meeting provided a venue for experts to further clarify their positions on definitional statements, advocate for their particular viewpoint, and discuss revised language in real-time. In all rounds of voting, the focus of the vote was on a series of statements or phrases within the overall definition that were under consideration either for their scientific merit or for their specific phraseology. For voting on these definitional statements, a threshold of 80% agreement was considered as “consensus” among the experts. This threshold was chosen as an appropriate cut-off based on previous examples of the Delphi method (Jorm, 2015; Eubank et al., 2016; Stern et al., 2018) and literature that suggested at least 80% agreement is needed to achieve content validity in a group of 10 or more experts (Lynn, 1986).

**Figure 1:**
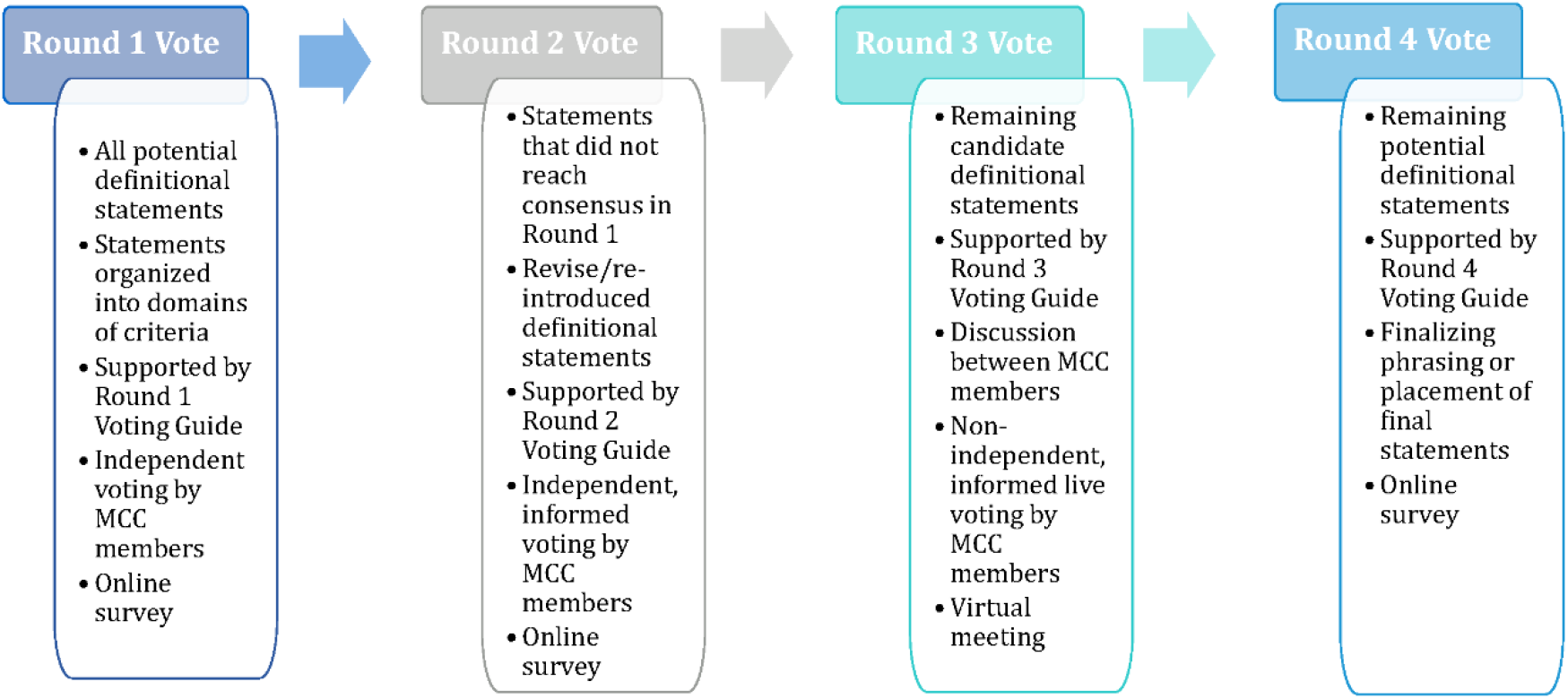
A modified Delphi process was employed to develop a misophonia consensus definition. In four rounds of voting, a Misophonia Consensus Committee (MCC), comprised of subject-matter experts, evaluated potential definitional statements about misophonia. Each round of voting differed in its intended purpose, what information the Committee relied on to make its determinations, and/or the format of voting.

## Materials and Methods

### Define the Project Objective and Identify Consensus Method

We first defined the overall objective of the consensus project: *to identify and publish a consensus definition of misophonia for the scientific community*. This objective served as an anchor point for experts who participated in the project and grounded the consensus process to its original goal throughout the project. The consensus process incorporated a modified Delphi method (Gustafson et al., 1973) and took place between June 2020 – January 2021. Staff from the Center for Strategic Philanthropy served as the facilitators for the consensus process and are referred to hereafter as the “facilitating team” or “facilitator.” A member of the MRF Scientific Advisory Board was included on the expert committee who developed the consensus definition to participate in the Delphi method process and serve as a liaison to the MRF Board.

### Establish Expert Committee

The consensus definition process required interdisciplinary input and participation from clinicians and researchers with diverse expertise, varied professional experiences, and knowledge of misophonia. Although there is little firm guidance on the ideal size of a Delphi expert panel (Jorm, 2015), findings from larger panels (e.g., more than 10) tend to be more stable than those from smaller panels as individual responses within larger groups have less of an influence over the ultimate outcome. A 15-person Misophonia Consensus Committee (MCC) was assembled throughout August – September 2020 to serve as the expert panel. Fifteen Committee members represented an ideal balance between stable responses (i.e. three opinions could diverge from the majority to still reach the pre-set consensus threshold of at least 80%) and administrative feasibility.

Potential members were identified as those formerly or currently engaged with the MRF through participation in convenings, engagement in grant review, service on the MRF Scientific Advisory Board, or as funded investigators. MCC members were also identified through recommendations from current MRF Board members or through independent research conducted by the facilitating team. Members of the Committee had diverse experiences in fields related to misophonia (audiology, neuroscience, psychology, neuropsychology, and psychiatry); expertise in clinical practice, development of definitions, diagnostic criteria, or measurement tools; and represented a range of career stages, geographies, nationalities, and genders.

As Committee members were recruited and onboarded, they were informed about: the overall objective of the project; the modified Delphi process and the anticipated timeline; guiding principles that Committee members were asked to commit to, including collaboration, objectivity, open-mindedness, and transparency; and authorship attribution and credit. Committee members were also required to agree to statements regarding conflicts of interest and confidentiality.

The MCC first convened via virtual meeting at the end of September 2020 to meet each other and gain additional familiarity with the facilitating team and the consensus process. The first round of voting launched in early October 2020. Round 2 ran from late November – early December 2020, and the Round 3 voting meeting was held in early January 2021. A fourth and final round of voting was used to finalize the definition by mid-January 2021. All 15 members participated in Rounds 1 and 2 of voting. In Round 3, 14 members participated in the first seven votes and 13 members participated in votes 8-19. Round 4 involved the participation of 14 members. “Consensus” was considered as 80% agreement of all Committee members present when a given vote was conducted.

### Systematic Literature Review

Committees who use Delphi methods may adopt different approaches to conduct systematic literature reviews. For example, some applications of the method will first establish the expert panel and then task the same committee to source the literature that they and their peers will evaluate during the consensus process (Venkatesan et al., 2019). Here, we elected to streamline this process by having the facilitator identify references at the same time as the Misophonia Consensus Committee was assembling. All identified references were then presented to the Committee for their consideration in the first round of evaluation and voting. Importantly, MCC members could identify additional references to supplement those identified by the facilitator, if necessary.

Delphi methods may also include an initial step whereby select members of the Committee first evaluate the level of evidence in each reference and thus categorize the “quality” of each potential statement under consideration (Eubank et al., 2016); these levels could range from randomized controlled trials (considered to be the highest level of evidence) to expert opinions (lowest level). However, rather than engage a select few MCC members to make these determinations for their colleagues, all MCC members received the same information regarding the literature, including type of publication, study design, and participant selection. This approach allowed the Committee to objectively evaluate the level of evidence for themselves as they considered and voted on candidate definitional statements.

References were sourced from *PubMed* and *Google Scholar*, as well as on the three preprint services, *PsyArXi*v, *bioRxiv* and *medRxiv*. References were identified as those published in English from 2001-September 2020 and that included “misophonia” in titles, keywords, and/or abstracts. References were also identified from citations in papers sourced by these criteria.

### Identifying Definitional Statements

Within each reference, we identified the specific language that authors used to define, describe, or characterize misophonia. This language was often located in the abstract and introduction of the publication. In other cases, misophonia was described in the results or conclusion, as the purpose of the publication was to report the outcomes of research focused on characterizing misophonia symptoms or other features. The sentences and statements that described or defined misophonia were extracted verbatim from each reference.

From the systematic literature review, we assembled a Microsoft Excel database of all definitional statements that had been extracted from the original sources in as close to the original wording as possible. Next, we identified common themes within the definitions, which we identified as Primary Domains of Criteria, and categorized the statements according to these domains.

### Developing Survey Questions and Fielding Surveys

The definitional statements identified during the literature review were further analyzed to derive concepts that could be written into survey questions. We continued working within the Excel database to classify these statements according to increasing levels of detail, including specific words or phrases and the frequency with which they appeared in the literature. From this database, we developed a detailed outline of definitional statements that served as the structure for the subsequent surveys and content of survey questions.

SurveyMonkey was used to manage Rounds 1, 2, and 4 of voting; Round 3 included discussion and polling via Zoom. Survey questions were written as short, declarative statements about a single concept that a Committee member could indicate their agreement or disagreement with. Although there are multiple ways to write Delphi process survey questions (Jorm, 2015), we aimed to minimize the number of choices presented to the MCC about each concept. This approach was selected over others (such as those that use a Likert scale) to ensure that statements could move through the consensus voting process more efficiently with fewer opportunities to “divide the vote.” In all surveys, the MCC had the opportunity to provide comments about the questions, propose alternative phrasing, or indicate concepts that may not have been included in the survey questions but should be considered. The response options varied depending on the round of voting (see below).

While it is not required for the Delphi process, some Delphi studies provide the expert panel with additional information to inform their decisions. Here, the Misophonia Consensus Committee received a comprehensive voting guide for each round of voting that included information specifically relevant to that round.

### Developing Points of Consensus Using a Modified Delphi Process

#### Round 1

In Round 1, the MCC evaluated the definitional statements presented in the Round 1 Survey questions based on their expertise and the results of the literature review that were presented in a companion Round 1 Voting Guide. For each Round 1 question, the survey included three response options as well as an open-text comment box where the Committee could explain their thought processes, offer evidence or citations, or propose alternative wording even if they agreed with the premise of the statement.

In Round 1, the most common answer options included:

- **Agree**: selected if the statement should be included in the consensus definition, based on the available scientific evidence;
- **Disagree**: selected if the statement should not be included in the consensus definition, as written, based on the available scientific evidence; or
- **Insufficient Information**: selected if there was insufficient evidence to determine whether or not the statement should be included in the consensus definition.

On some questions, the Committee was asked whether a specific feature or characteristic was considered to be essential to misophonia or whether it varied in its occurrence. For these types of questions, the answer options were “Always,” “Sometimes,” or “Insufficient Information” with the open-text box option available as well.

A Round 1 Voting Guide accompanied the Round 1 Survey and included detailed information about the references identified in the literature review, including the original wording of definitional statements extracted from each reference. Both the survey and the voting guide – including the references − were organized by Primary Domain of Criteria. Because survey questions were often synthesized from definitional language that appeared in multiple references, it was not feasible to identify unique references for each individual survey question. However, references were identified for each Primary Domain and sub-themes for the Committee to refer to as they evaluated statements related to a broad definitional concept (such as auditory stimuli that may trigger symptoms of misophonia).

The 15 MCC members had three weeks to complete the Round 1 Survey. After three weeks, the response frequencies for each question were analyzed and the feedback provided in the Round 1 Survey comments was evaluated. An 80% agreement threshold (12 of 15 MCC members) was considered as consensus to either include the statement in the final definition, or exclude the statement from further consideration. Statements that did not meet consensus in Round 1 were re-evaluated in Round 2.

#### Round 2

In Round 2, the Committee re-evaluated the definitional statements that did not reach consensus in Round 1. The Committee based their Round 2 evaluation on their expertise, the results of the literature review, *and* the aggregated results and anonymized comments from Round 1 that were provided in a Round 2 Voting Guide. For most Round 2 questions, a question from Round 1 was revised based on MCC comments and presented in the Round 2 Survey as a choice between the original language that reached partial agreement and the revision. A third option – “None of the above/Insufficient evidence to include in the definition” – was also presented, as well as an open-text comment box. In other cases, multiple questions from Round 1 were condensed into a single multiple-choice question in Round 2.

The Round 2 Survey included three different formats of questions and responses that depended on the information under evaluation:

- **Example Question 1:** Please select the *one* option that you *most* agree with:
  - Example responses:
    ▪ All original statements from Round 1
    ▪ None of the above/Insufficient evidence to include in the definition
- **Example Question 2**: Please select the *one* option that you *most* agree with:
  - Example responses:
    ▪ Original statement(s) from Round 1
    ▪ Revised statement(s) that incorporated MCC feedback from Round 1
    ▪ None of the above/Insufficient evidence to include in the definition
- **Example Question 3:** Please select the option(s) that you *most* agree with. You may select more than one option if you agree with them; however, if you feel that none fit, please select “none of the above.”
  - Example responses:
    ▪ All original statements from Round 1
    ▪ None of the above/Insufficient evidence to include in the definition

A Round 2 Voting Guide accompanied the survey and included information that the MCC used to evaluate Round 2 questions, including:

- Context for a batch of Round 2 questions and response options – the same information was available in the Round 2 Survey
- The Round 1 statement(s)/question(s) that contributed to a given Round 2 question
- Aggregated results for the relevant Round 1 question(s)
- Anonymized comments from MCC members on the relevant Round 1 question(s)
- Relevant references from the literature review for the Round 2 question

Voting guides were individually customized for each MCC member to indicate their votes and comments on the relevant Round 1 question(s).

The MCC again had three weeks to complete the survey. Response frequencies for each question were analyzed and the feedback provided in survey comments was reviewed. An 80% agreement threshold (12 of 15 MCC members) was considered as consensus to either include the statement in the definition or exclude the statement from further consideration. Select statements that did not meet consensus were re-evaluated in Round 3.

#### Round 3

By the conclusion of Rounds 1 and 2, the Committee had reached consensus on a sufficient number of statements and a draft of the definition was developed. At this point, statements that had met consensus to include in the definition were synthesized and written into prose for MCC review and feedback. Prior to the third round of voting, the MCC was provided with a Round 3 Voting Guide that included two drafts of the definition:

- Version 1 incorporated all statements that met consensus in Rounds 1 and 2;
- Version 2 included the same information as in Version 1 but with the addition and identification of statements that would be discussed and voted on in Round 3.

The statements identified for discussion and voting in Round 3 were selected because they were either close to reaching consensus in Round 2 (one or two votes shy) and/or MCC feedback indicated that they were integral or helpfully additive to the definition (e.g., examples of statements that met consensus to include in the definition).

The third round of voting was held in early January 2021 in a 2-hour virtual meeting. Thirteen of the 15 MCC members voted on all statements with a fourteenth member present for the first seven votes. The statements were considered one at a time and presented via PowerPoint slide with the surrounding paragraphs in which they were found in Version 2 of the draft definition. This approach allowed the Committee to evaluate each statement in context.

Prior to any discussion, a proposed definitional statement was presented, and the MCC voted via poll questions: “Yes” in support of its inclusion as presented in Version 2 of the definition and on the slide; or “No” to indicate further discussion or exclusion. If greater than 80% consensus was reached on this first vote, the floor was briefly held open for discussion before the statement was considered as “accepted” and the Committee moved to the next statement. If the first vote yielded less than 80% consensus, then the statement was discussed, potentially revised in real-time, and a second vote was held.

There were multiple outcomes for statements in the Round 3 vote:

Included in the final definition exactly as it was presented in Version 2 of the definition and discussed during the Round 3 meeting;

- Included after the language was revised based on Round 3 discussion;
- Included in principle with the MCC to revisit the phrasing, the statement’s location in the definition, or its integration with other parts of the definition in the next revision (Version 3) of the definition;
- Revised in Version 3 of the definition because the statement had MCC support but no consensus in Round 3 *and* the MCC agreed to revisit it;
- Excluded based on consensus reached by the MCC to exclude; or
- Excluded based on no consensus reached in Round 3 and a lack of MCC support to continue considering the statement.

#### Round 4

Although a 3-round Delphi process was initially planned, we elected to hold a fourth round of voting to finalize language on six statements that had MCC support but no final decision after the Round 3 meeting. The Round 4 Survey was managed through SurveyMonkey and accompanied by a Round 4 Voting Guide that reflected the discussion and vote outcomes from Round 3. This Round 4 Voting Guide also tracked how the statements that met consensus in Round 3 were incorporated into the revised draft definition (Version 3). In the 6-question Round 4 Survey, MCC members were presented with two answer choices that would determine the location of a concept in the definition (either Location A or B) or indicate their agreement/disagreement with specific phrasing. Feedback and/or proposed revisions were also encouraged via a comment box. The results from Round 4 were incorporated into the draft definition to arrive at the final version of the definition – Version 4.

## Results

### Systematic Literature Review

Sixty-eight references were identified during the literature review as meeting the pre-established criteria (described in the *Methods*) and that included a description, definition, or characterization of misophonia (Table 1).

**Table 1:**
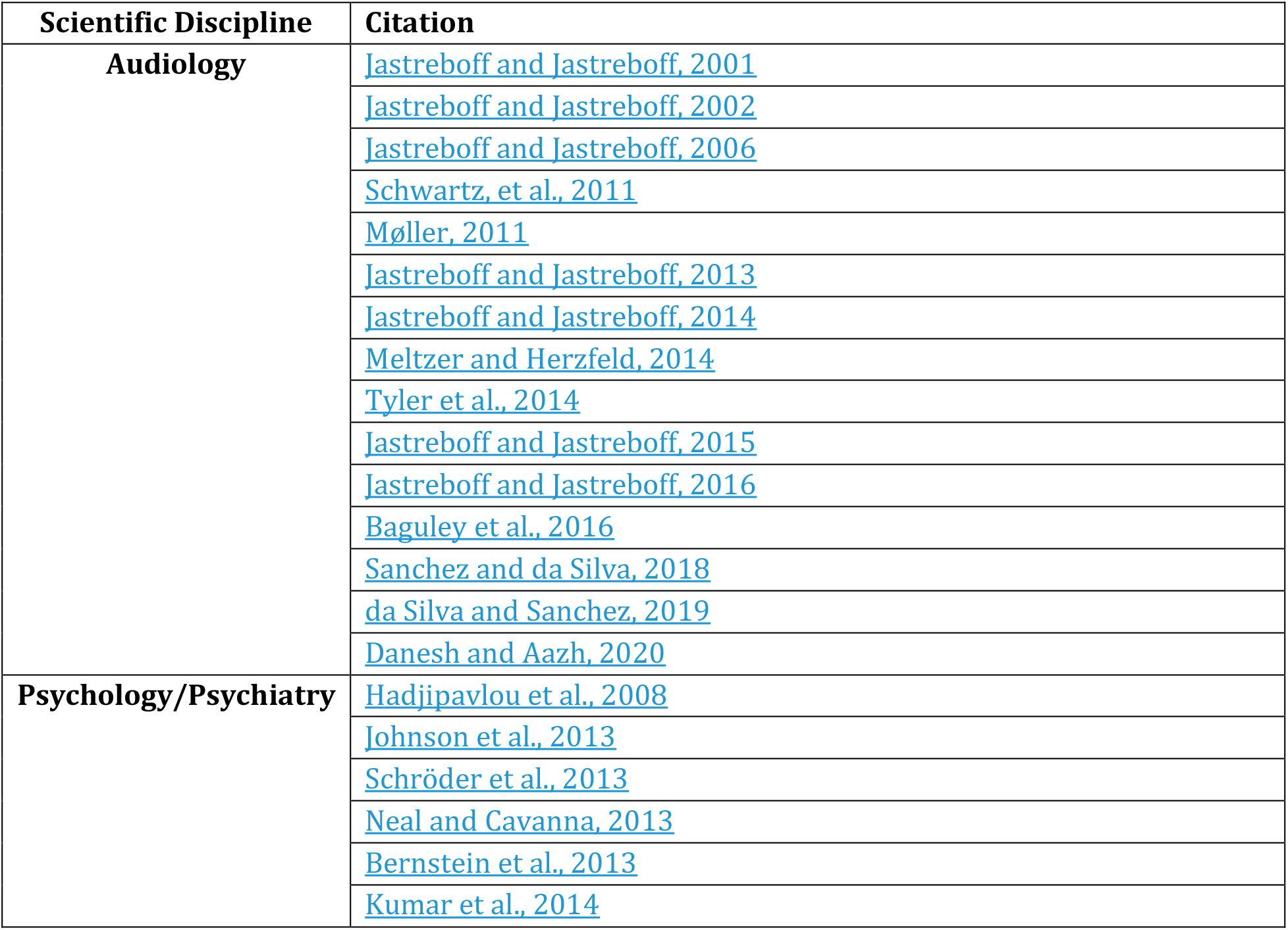
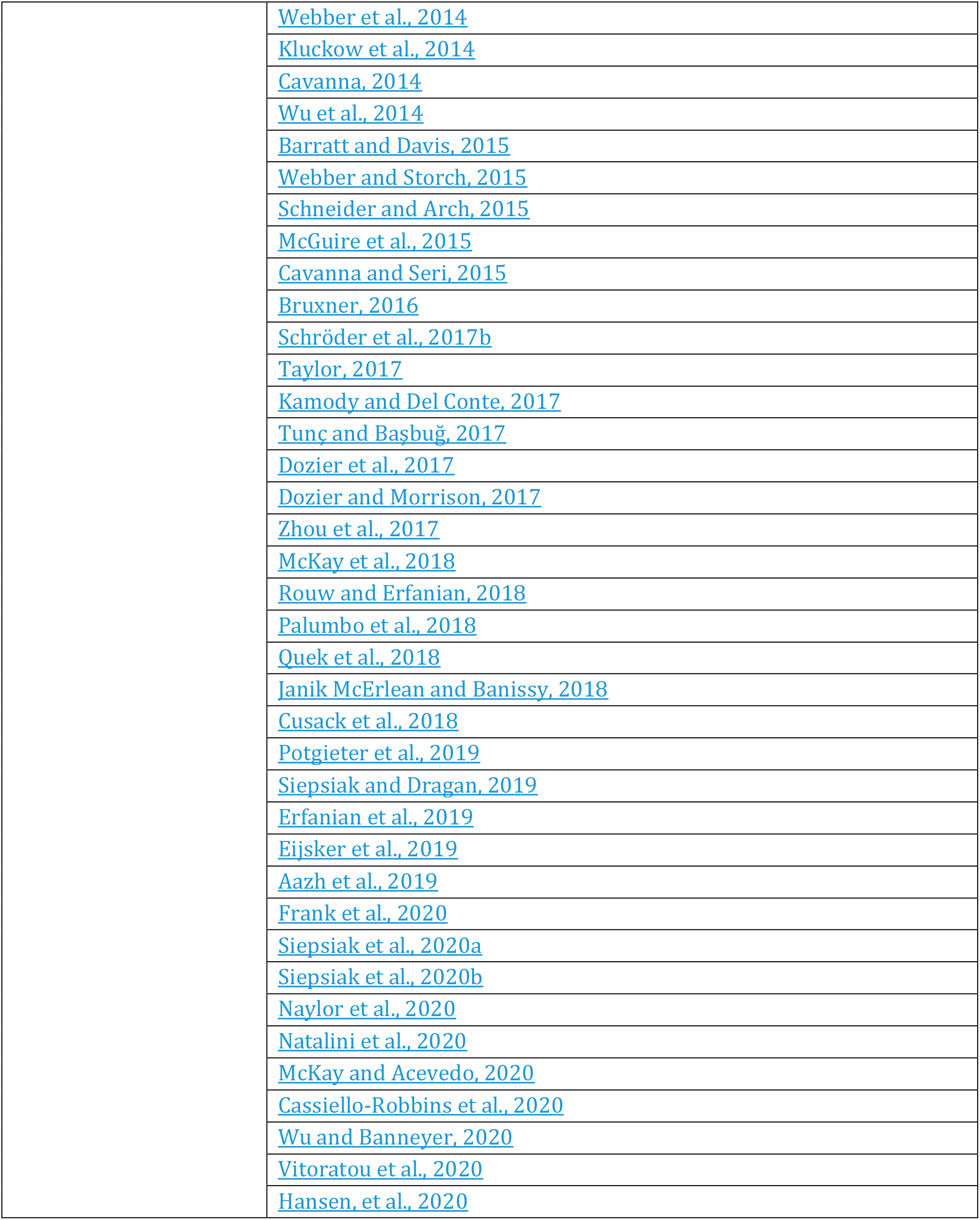
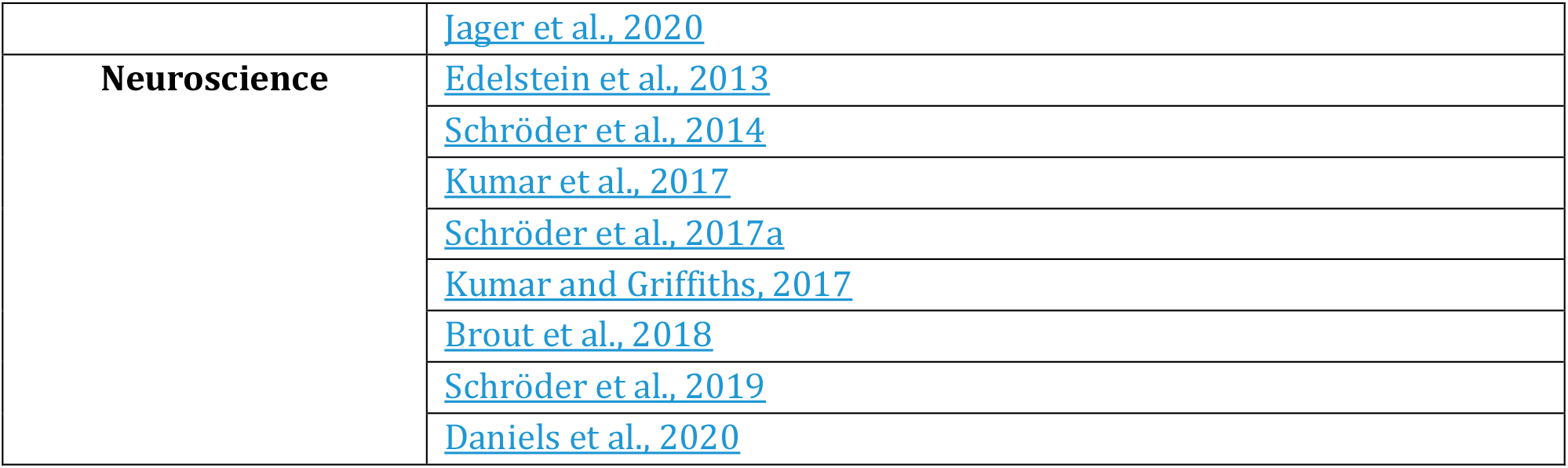
68 references that included definitional statements about misophonia were identified through a systematic literature review. References were sourced from *PubMed* and *Google Scholar*, as well as on the three preprint services, *PsyArXi*v, *bioRxiv* and *medRxiv*. References were identified as those published in English from 2001-September 2020 and that included “misophonia” in titles, keywords, and/or abstracts. References were also identified from citations in papers sourced by these criteria. Candidate definitional statements were sourced from all 68 references. References are organized in Table 1 according to their scientific discipline.

From each reference, definitional statements about misophonia, as well as other key information, were extracted and shared with the Committee (Table 2). Committee members referred to this information to evaluate the strength of the scientific evidence that supported candidate definitional statements about misophonia.

**Table 2:** Key information extracted from a systematic review of the misophonia literature. From each reference identified during the systematic literature review, multiple pieces of information were extracted and presented to the Misophonia Consensus Committee to inform the misophonia definition development process.

| Type of Key Information | Specific Information | Examples |
| --- | --- | --- |
| Bibliographic Information | Full citation |  |
|  | Publication DOI/Link |  |
|  | PDF of reference |  |
| Classification Information | Scientific discipline of references | Audiology |
|  |  | Neuroscience |
|  |  | Psychiatry/Psychology |
| Description of References | Type of reference or study | Peer-reviewed observational study |
|  |  | Peer-reviewed interventional study |
|  |  | Peer-reviewed review article |
|  |  | Peer-reviewed case report |
|  |  | Textbook chapter |
|  |  | Non-peer reviewed article (e.g. in professional newsletter, on website) |
|  |  | Non-peer reviewed observational clinical study (i.e. preprint manuscript) |
|  |  | Non-peer reviewed case report |
|  |  | Scientific poster abstract |
|  |  | Editorial |
|  |  | Commentary |
| Detailed Information from Reference | Study participants (not always described) | Number of study participants |
|  |  | Characteristics of participants – in experimental and control groups |
|  |  | Recruitment methods |
| Definitional statements | Identified and extracted verbatim from each reference |  |

### Identifying Definitional Statements

The Excel database built from the definitional statements extracted from 68 references included 551 individual statements. Statements were first extracted from the original sources as close to the original wording as possible, such as: “*Misophonia is a chronic condition in which specific sounds provide intense emotional experiences and autonomic arousal within an individual*” (Cusack et al., 2018). Next, common themes were identified within the definitions, such as language that generally described misophonia, or more detailed descriptions of the emotional or physiological reactions that may be evoked by trigger stimuli. Twelve such themes, or “Primary Domains of Criteria,” were identified from the literature (Table 3).

**Table 3:** Twelve primary domains of criteria about misophonia were identified during the literature review. Twelve thematic areas about misophonia emerged within all of the definitional statements that were identified in the published literature.

| Primary Domain | Description |
| --- | --- |
| Domain 1: General Description | Fundamental information that would be found in the first statements of the definition, such as whether misophonia is a condition or disorder, its potential spectrum nature, and how it can be briefly described. |
| Domain 2: Trigger Stimuli | General statements about misophonic triggers, what types of stimuli modalities they tend to be, examples, and common features. |
| Domain 3: Emotional Reactions | General statements about emotional responses to trigger stimuli, all negative emotions, specific emotions related to |
|  | anger or anxiety, words to describe emotions (e.g. strong, extreme), timescale and transition of reactions. |
| Domain 4: Physiological Reactions | General statements about physiological responses to triggers, specific reactions, and descriptors (e.g. sudden, extreme). |
| Domain 5: Behavioral Reactions | General statements about behavioral responses to triggers or in anticipation of them, descriptors, transitions between behaviors, and targets of these reactions (e.g. person or object). |
| Domain 6: Attentional Reactions | Examples such as hyper-focus or obsession. |
| Domain 7: Influences on Reactions | Description of physical characteristics of stimuli (e.g. pitch/frequency) and whether these play a role, context of stimuli, and individual variables. |
| Domain 8: Insight & Awareness | Language regarding whether people have insight into and awareness of their reactions, as compared to other people, as well as increased awareness of trigger stimuli compared to other stimuli. |
| Domain 9: Functional Impairment | General descriptions of potential impairments, examples of occupational/academic or social impairments. |
| Domain 10: Coping Strategies | Example approaches that may be employed to cope with distress caused by triggers. |
| Domain 11: Onset and Course | Age of onset for misophonia, and language about the potentially chronic nature of the disorder as well as potential familial links. |
| Domain 12: Misophonia is Not Otherwise Explained By | Description of auditory functioning in individuals with misophonia, consideration of auditory perception conditions, medical conditions, and psychiatric conditions. |

Statements were then categorized within the 12 Primary Domains. In some cases, the definitional sentence or statement, as originally written in the reference, was clearly aligned with only one Primary Domain. For example, the statement *“[Misophonia] includes a broad spectrum of emotions including but not limited to fear,”* (Jastreboff and Jastreboff, 2002) was assigned to Domain 4: Emotional Reactions. In other cases, the original definitional sentence from the reference covered multiple domains and was thus divided into multiple distinct statements and Primary Domains. For example, the definitional sentence “*Those with misophonic symptoms often experience significant impairment across occupational/academic, familiar/home-based and social functioning in response to the disgust, anger, and distress caused by auditory cues,”* (Webber and Storch, 2015) was categorized as:

- Domain 9: Functional Impairment – “*Those with misophonic symptoms often experience significant impairment across occupational/academic, familiar/home-based and social functioning…”*
- Domain 4: Emotional Reactions – “…*in response to the disgust, anger, and distress caused by…*.*”*
- Domain 2: Triggering Stimuli – “*…auditory cues*.*”*

### Developing Survey Questions

The 551 individual definitional statements were further analyzed to identify additional levels of detail. These sub-themes were used to assemble a detailed outline of all potential statements that were then used to develop survey questions. For example:

- Primary Domain: Trigger Stimuli
  - Secondary Theme: Auditory Triggers
    ▪ Tertiary Theme: Produced by the Human Body
      - Example: Chewing

This classification method was used to further resolve the detail within definition statements as well as identify specific language to be incorporated into the survey questions. This approach ensured that the survey questions accurately reflected the content of the definitional statements that were extracted from the misophonia literature.

The first round of survey questions presented short, declarative statements about a single concept, such as: “*Misophonia trigger stimuli are repetitive*.*”* Subsequent rounds of voting included questions that qualified these concepts, using terms such as “*may*,” “*usually*,’ or “*often*,” and presented increasingly complex statements or sentences to the MCC as they refined the language and location of statements within the overall definition.

### Developing Points of Consensus Using a Modified Delphi Process

#### Round 1

The Round 1 Survey included 199 questions that covered all 551 potential definitional statements identified in the systematic literature review. The survey covered 31 pages and was organized by the 12 Primary Domains or Criteria with secondary domains identified, when appropriate. Statements met consensus at 80% or more agreement (12/15 MCC members) to either include in the definition or exclude from further consideration. The results of Round 1 are illustrated in Figure 2.

**Figure 2:**
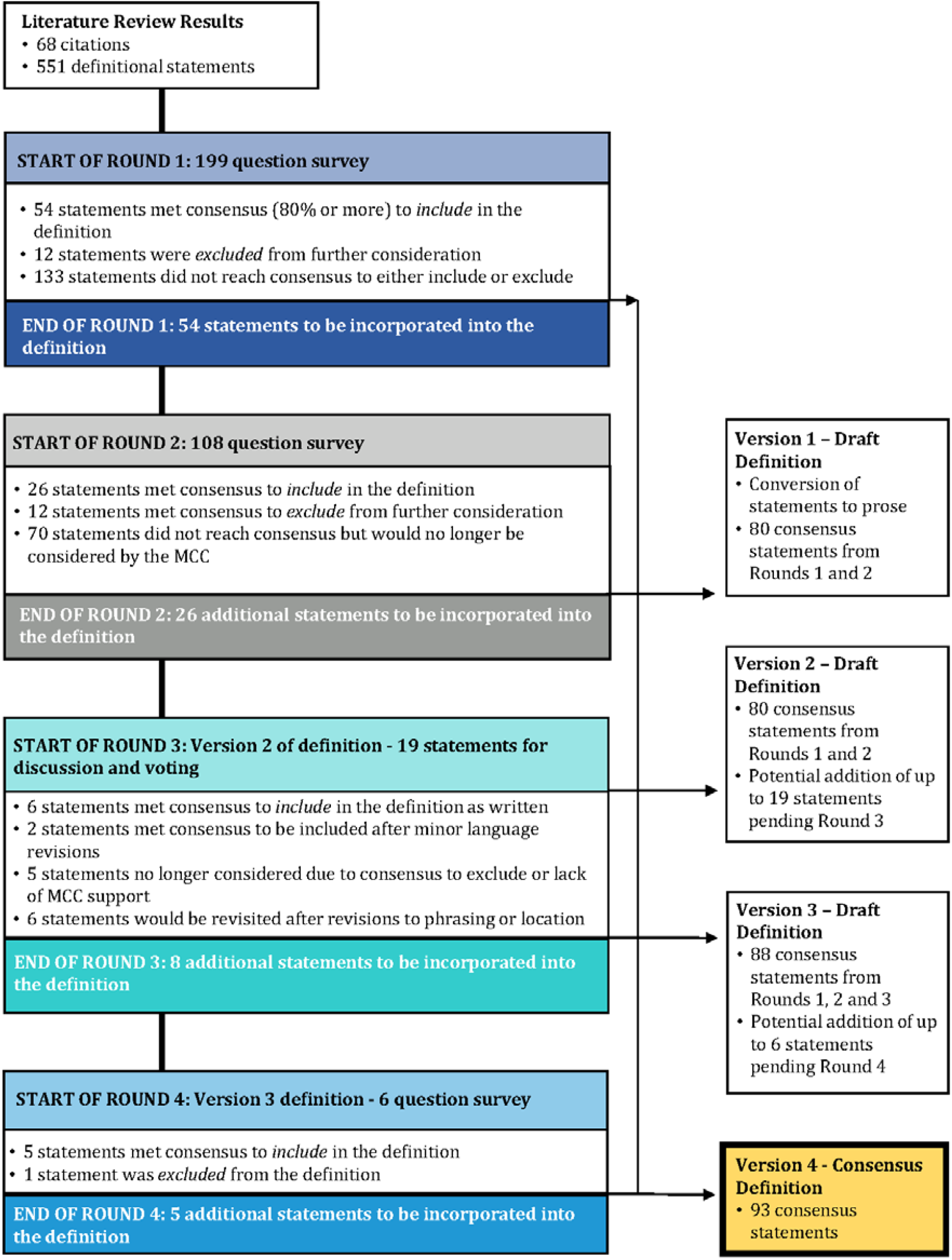
Methodology and results of a modified Delphi method to develop a consensus definition of misophonia. Through four rounds of evaluation and voting on potential definitional statements that were extracted from the published scientific literature, a committee of experts developed a consensus definition of misophonia.

Fifty-four statements met consensus in Round 1 to include in the definition by at least 80% of MCC members selecting the response option “Agree” or indicating that the statement was at least “Sometimes” seen in misophonia. These 54 statements covered 10 of the 12 Primary Domains of Criteria. While the Committee agreed to include these statements in the final definition, members provided minor feedback that was later incorporated as the first version of the definition was drafted.

Twelve statements were excluded from further consideration after Round 1 after having met one of three conditions:

- At least 80% of MCC members selected the response options “Disagree” or “Insufficient Information;”
- No MCC members agreed with the original statement (i.e. 0% “Agree”) with remaining responses split between the “Disagree” and “Insufficient Information” responses options. Comments from the Committee indicated that there was no support for the concept and that it was not worthwhile to reevaluate in Round 2.
- A minority of MCC members (three or fewer) agreed with the statement while a related or companion statement, such as one that presented the opposite concept or the same concept with different phrasing, reached consensus to include in the definition.

Statements that did not meet consensus in Round 1 were re-evaluated in Round 2; 133 statements met these criteria and MCC feedback on these statements was incorporated in revisions for the MCC to evaluate in a Round 2 Survey.

#### Round 2

The Round 2 Survey included 108 questions that were based on the 133 statements that did not meet consensus in Round 1. The survey covered 37 pages and was again organized by Primary Domain of Criteria with each survey page including context to frame the specific batch of questions under consideration. Statements again met consensus at 80% or more agreement (12/15 MCC members) to either include in the definition or exclude from further consideration. The results of Round 2 are illustrated in Figure 2.

Twenty-six statements met consensus in Round 2 to include in the definition and represented 9 of the 12 Primary Domains of Criteria. These 26 statements were combined with the 54 statements that met consensus in Round 1 for a total of 80 statements that met consensus to include in the definition after two rounds of voting. As in Round 1, MCC members provided feedback in Round 2 on statements that they thought should be included in the definition; this feedback was incorporated as the first version of the definition was drafted.

Twelve statements met consensus in Round 2 with 80% or more MCC agreement to exclude from the definition. Seventy statements did not reach consensus in Round 2 to either include or exclude from the definition. The MCC’s responses and feedback on these 70 statements was carefully evaluated and, to ensure the best use of the Committee’s effort in subsequent rounds of voting, 52 of these 70 statements were excluded from further consideration because they:

- had support from less than two-thirds of the Committee after two rounds of voting and MCC-suggested revisions; and/or
- were not considered to be integral to the final definition, based on MCC comments; and/or
- were redundant to other statements that had met consensus to either include in or exclude from the definition.

Nineteen of the 70 statements that did not reach at least 80% consensus in Round 2 were specifically identified for Round 3 discussion and voting because they:

- were two or fewer votes shy of reaching consensus in Round 2; and/or
- MCC feedback on these and other statements indicated that they were integral or helpfully additive to the definition (such as by serving as examples of statements that are included in the definition).

One of the Round 2 questions concerning emotional reactions included multiple response options that met consensus to include in the definition as well as one response that did not meet consensus but was considered to be worthy of discussion in Round 3. Therefore, this statement (Round 2, Question 39) counted as both one of the 26 statements to include in the definition after Round 2 as well as one of the 19 statements that would be discussed in Round 3.

#### Round 3 and Draft Versions 1 and 2 of the Misophonia Definition

By the conclusion of Rounds 1 and 2, 80 statements had reached consensus and a draft definition – Version 1 – was developed that incorporated these 80 statements and the feedback that the MCC provided on them in Rounds 1 and 2. A second definition draft – Version 2 – was simultaneously drafted that reflected all 80 consensus statements as well as the 19 statements that were pending discussion and voting in Round 3. The MCC was provided with both Versions 1 and 2 of the definition in their Round 3 Voting Guide to demonstrate that they had already reached consensus on a definition but that they may elect (or not) to supplement that definition with statements that they would consider in Round 3. The results of Round 3 are illustrated in Figure 2.

During the Round 3 meeting, held via Zoom, the MCC discussed and voted on 19 statements. These 19 statements were close to reaching consensus in Rounds 1 or 2 and/or the MCC’s comments indicated were important to the final definition. There were multiple outcomes for statements in the Round 3 vote:

- six statements: included in the final definition exactly as they were presented in Version 2 of the definition/during the Round 3 meeting;
- two statements: included after the language was revised based on Round 3 discussion;
- four statements: included in principle with the MCC to revisit the phrasing, the statements’ location in the definition, or their integration with other parts of the definition the next revision (Version 3) of the definition;
- two statements: revised in Version 3 of the definition with the MCC to revisit the revised language because the statements had MCC support but did not reach consensus in Round 3;
- one statement: excluded based on consensus reached by the MCC to exclude; and
- four statements: excluded based on no consensus reached in Round 3 and a lack of MCC support to continue considering the statements.

#### Round 4 and Draft Version 3 of the Misophonia Definition

After Round 3, 8 additional statements were incorporated into the misophonia definition to develop the next draft – Version 3. Six statements were identified during the Round 3 discussion as warranting follow-up consideration from the MCC to determine final phrasing or location in the definition; these six statements were evaluated in a Round 4 Survey. Any revisions that arose from the Round 4 Survey would be incorporated into the next draft of the definition – Version 4.

Fourteen MCC members voted on these six statements in the Round 4 Survey. Because the MCC had reached 80% or more agreement in Round 3 to include four of these six statements in the definition, a simple majority (50% or more) in Round 4 determined the outcome of these statements. The other two statements assessed in Round 4 had not yet reached consensus in Round 3 and thus the 80% threshold still applied.

The MCC’s Round 4 voting results surpassed the required thresholds for all six statements (i.e. 50% for four statements and 80% for the remaining two). However, comments from multiple MCC members on one of the six statements indicated that the concept was still confusing and may not be important for the definition. Therefore, although more than 50% of the MCC agreed with including this statement in the definition, the totality of feedback that the MCC shared in both the Round 3 discussion and on the Round 4 survey led to the conclusion that this specific statement should be eliminated from the definition.

After Round 4, 5 additional statements were integrated into the final draft of the definition – Version 4. This fourth and final version of the draft definition incorporates 93 individual definitional statements that have all met 80% or greater Committee consensus. The results of Round 4 are illustrated in Figure 2.

### Consensus Definition of Misophonia

#### GENERAL DESCRIPTION

Misophonia is a disorder of decreased tolerance to specific sounds or stimuli associated with such sounds. These stimuli, known as “triggers,” are experienced as unpleasant or distressing and tend to evoke strong negative emotional, physiological, and behavioral responses that are not seen in most other people. Misophonic responses do not seem to be elicited by the loudness of auditory stimuli, but rather by the specific pattern or meaning to an individual. Trigger stimuli are often repetitive and primarily, but not exclusively, include stimuli generated by another individual, especially those produced by the human body. Once a trigger stimulus is detected, individuals with misophonia may have difficulty distracting themselves from the stimulus and may experience suffering, distress, and/or impairment in social, occupational, or academic functioning. The expression of misophonic symptoms varies, as does the severity, which ranges from mild to severe impairments. Some individuals with misophonia are aware that their reactions to misophonic trigger stimuli are disproportionate to the circumstances. Misophonia symptoms are typically first observed in childhood or early adolescence.

#### REACTIONS TO MISOPHONIC TRIGGERS

In response to specific trigger stimuli, individuals with misophonia may experience a range of negative affective reactions. Anger, irritation, disgust, and anxiety are most common, though some individuals may experience rage. Misophonic triggers may evoke increased autonomic arousal such as increased muscular tension, increased heart rate, and sweating.

Trigger stimuli may also evoke strong behavioral reactions such as agitation or aggression directed towards the individual producing the stimulus. On rare occasions, aggression may be expressed as verbal or physical outbursts although these responses are seen more in children with misophonia than in adults. Individuals with misophonia often engage in behaviors to mitigate their reactions to triggers such as: avoiding or escaping from situations in which they encounter trigger stimuli; seeking to discontinue the triggering stimuli; mimicking or reproducing the triggers.

#### INFLUENCES ON REACTIONS

The strength of an individual’s reaction to a misophonic trigger stimulus may be influenced by multiple factors including but not limited to: the context in which the stimulus is encountered; the individual’s perceived degree of control over the stimulus source; and the interpersonal relationship between the individual with misophonia and the source of the trigger. Self-generated stimuli typically do not evoke the same aversive responses as stimuli produced by other people.

#### FUNCTIONAL IMPAIRMENTS

Individuals’ reactions to misophonia triggers may cause significant distress, interfere with day-to-day life, and may contribute to mental health problems. Individuals with misophonia may experience functional impairments that range from mild to severe including but not limited to impaired occupational and/or academic functioning, concentration difficulties, and an inability to perform important work tasks. Individuals may also experience impaired social functioning, strained social relationships, and social isolation resulting from their misophonia symptoms.

#### RELATIONSHIP TO OTHER CONDITIONS/DISORDERS

Misophonia can be present in people with or without normal hearing thresholds, and can occur alone or with the auditory conditions of tinnitus and hyperacusis. Misophonia can also occur with neurological or psychiatric conditions or disorders including but not limited to: anxiety disorders, mood disorders, personality disorders, obsessive compulsive related disorders, post-traumatic stress disorder, autism spectrum disorder, and attention deficit hyperactivity disorder. For any given individual, the symptoms of misophonia should not be better explained by any co-occurring disorders.

#### MISOPHONIC TRIGGERS

Although each person may have their own pattern of triggers, some stimuli serve as common misophonic triggers. Auditory triggers are most common, although individuals with misophonia may also identify distress in response to visual triggers.

Sounds associated with oral functions are among the most often reported misophonic trigger stimuli, such as chewing, eating, smacking lips, slurping, coughing, throat clearing, and swallowing. Nasal sounds, such as breathing and sniffing, often serve as triggers as well. Auditory triggers may also include non-oral/nasal sounds produced by people such as pen clicking, keyboard typing, finger or foot tapping and shuffling footsteps, as well as sounds produced by objects, such as a clock ticking, or sounds generated by animals. Visual triggers have been reported to include stimuli such as cracking knuckles and jiggling or swinging legs, as well as visual stimuli associated with an auditory trigger, such as watching someone eat.

## Discussion

Misophonia was first named and described in 2001 (Jastreboff and Jastreboff, 2001; Jastreboff and Jastreboff, 2002) but has since been characterized and defined differently by researchers and clinicians from different fields and with varying areas of expertise. The lack of a common, foundational definition has made it difficult to compare study cohorts, evaluate treatment approaches, and validate tools to diagnose and assess the severity of misophonia. It is therefore essential that a common definition of misophonia be identified for individuals experiencing misophonia, the clinicians who support them, and researchers who seek to better understand this condition and evaluate treatments.

Here we present a consensus definition of misophonia that has been developed through a modified Delphi process by a 15-person committee of researchers and clinicians with diverse expertise and experiences related to misophonia. The definition reflects the outcome of four rounds of evaluation and voting by the Committee on definitional statements published in the misophonia scientific literature. The final, consensus definition incorporates 93 statements that each met consensus at 80% or more Committee agreement to include in the definition based on the currently available scientific and clinical evidence. This consensus definition drafted by the Misophonia Consensus Committee is intended to serve as a working definition for the field that can and should be validated, reevaluated, and revised as the research and clinical community’s understanding of misophonia evolves.

### Reflections on the Final Definition – Areas for Further Inquiry

The consensus definition incorporates nearly 100 statements. However, these represent a minority of all potential definitional statements that were extracted from the original literature review. The Misophonia Consensus Committee excluded concepts from the final definition because they agreed that the available scientific evidence was either inconclusive or explicitly did not support a concept or specific phraseology.

#### Broad Description of Misophonia

Misophonia has been broadly described in the literature as a condition (e.g. Edelstein et al., 2013; Johnson et al., 2013; Jager et al., 2020), syndrome (e.g. Cavanna and Seri, 2015; Taylor, 2017; Brout et al., 2018), or disorder (e.g. Schröder et al., 2013; Baguley et al., 2016; Kumar et al., 2017; Erfanian et al., 2019), and the Committee did not reach consensus until Round 4 to describe misophonia as a “disorder.” “Disorder” was ultimately determined to be a more accurate and useful descriptor than “condition” or “syndrome” for the purposes of the definition. The MCC felt that “disorder” correctly implicates the negative experience of individuals experiencing misophonia, can be useful in driving scientific inquiry to develop treatment models, and reinforces the professional and societal context around properly diagnosing, treating, and reimbursing care for misophonia. The Committee concluded that the scientific evidence regarding whether or not to classify misophonia as a “medical” (Cavanna and Seri, 2015) or “psychiatric” disorder (Schröder et al., 2013) is currently insufficient but that underlying organic etiology of the disorder cannot be ruled out. The Committee agreed that the available evidence did not support defining misophonia as a “reflex condition” (Dozier et al., 2017). Finally, although the name misophonia can be literally translated as “hatred of sound,” and is described this way in many publications, Committee members objected to including this translation in the definition as those with misophonia neither specifically feel hate nor do they necessary feel strong emotions only related to sound (i.e., some also have similar responses to visual triggers not associated with sounds, such as leg swinging).

#### Potential Mechanisms

The Committee agreed that the current literature did not yet support including language related to proposed biological, genetic, or behavioral mechanisms that may underlie misophonia. Whereas studies have postulated differential reactivity of different neural systems, such as those involved in emotional regulation, learning, and auditory processing (Jastreboff and Jastreboff, 2002; Jastreboff and Jastreboff, 2014; Schröder et al., 2017b), an understanding of the biological processes that underlie misophonia is currently under active investigation. The Committee concluded that *postulated* mechanisms do not belong in the definition at this time. Similarly, although a few case studies have identified multiple cases of misophonia within extended families (Cavanna, 2014; Sanchez and da Silva, 2018), the current available evidence does not support including language about a familial link to the disorder in the definition.

#### Prevalence, Onset, and Course

Multiple studies have estimated the prevalence of misophonia in different populations (Wu et al., 2014; Zhou et al., 2017; Quek et al., 2018; Rouw and Erfanian, 2018; Jager et al., 2020; Naylor et al., 2020; Siepsiak et al., 2020b) by using different diagnostic questionnaires and measurement tools (Jastreboff and Jastreboff, 2002; Schröder et al., 2013; Bernstein et al., 2013; Johnson et al., 2013; Wu et al., 2014; Rouw and Erfanian, 2018; Jager et al., 2020; Siepsiak et al., 2020a; Vitoratou et al., 2020). However, because these tools are based on different definitions for misophonia and most tools have not yet been psychometrically validated, the Committee agreed that it would be premature to include statements about the prevalence of misophonia in the consensus definition. Similarly, although the symptoms of misophonia are typically first observed/detected in childhood or early adolescence (Johnson et al., 2013; McGuire et al., 2015; Palumbo et al., 2018), the actual age of onset for the disorder is an area of active inquiry and the Committee determined that the consensus definition should not define the age of misophonia onset at this time. Finally, the Committee agreed that the available evidence does not yet support defining a “typical” course of misophonia over an individual’s lifetime – such as remaining stable or worsening – due to an absence of prospective and longitudinal studies.

#### Relationships to Other Conditions or Disorders

The Committee reached consensus to state that the symptoms of misophonia should not be better explained by auditory, psychological, and psychiatric disorders. However, Committee members felt that the etiology of misophonia and its relationships with other conditions are not yet clear and should not be included in the definition at this time. For example, the role of auditory functioning in misophonia is an area of active research and Committee members agreed that the definition should not include language regarding how misophonia specifically relates to hearing disorders. Similarly, ongoing research seeks to understand how misophonia relates to psychiatric disorders, as well as how misophonia may be influenced by psychological characteristics or individual personality factors. The field has not yet settled on these issues and Committee members agreed that it was not their role to make these determinations for the purposes of defining misophonia.

### Limitations

Methods to reach consensus within groups of experts may be influenced by the opinions of dominant individuals, coercion, or pressure to adopt certain opinions or viewpoints (Jorm, 2015). The Delphi method seeks to minimize these effects by maintaining independence and anonymity throughout multiple rounds of informed assessment and voting (Gustafson et al., 1973; Murphy et al., 1998). The method described here to develop a consensus definition of misophonia also included strong guards against groupthink by ensuring that MCC members represented multidisciplinary scientific and clinical backgrounds and had diverse expertise and training.

The Delphi method can be criticized for its adherence to anonymity early in the voting process which results in Committee members not fully benefiting from the expertise of their peers (Dalkey, 1969). We sought to balance the need for independent thought with informed assessment by sharing the anonymized results and comments of Committee members with each other after Rounds 1 and 2 of voting, as well as providing a “face-to-face” meeting in Round 3 when members could openly discuss the definition and advocate for their specific viewpoints (Gustafson et al., 1973).

Another potential limitation of the Delphi consensus method relates to the composition of the expert committee. The Delphi method does not provide formal guidance about who should be considered to be an expert for the purposes of selecting a consensus committee. In our study, we identified criteria for MCC member selection (see: *Methods*) during the initial planning stages of the project and then recruited members according to these criteria. More specifically, the MCC was comprised of individuals with professional clinical and research expertise that spanned audiology, auditory neuroscience, psychology, psychiatry, and cognitive neuroscience. The Committee did not include non-professionals or individuals who themselves suffer from symptoms of misophonia. Although the MCC was mindful of developing a definition that could be understood by a non-technical audience and is relevant for individuals experiencing misophonia, a committee comprised of other individuals with different expertise and experiences may have reached a different final definition.

To some extent, there is an unavoidable circularity inherent in developing a definition for misophonia using definitional statements from published research studies that have described individuals with misophonia in particular ways. Importantly, MCC member expertise was not restricted to misophonia per se, as members represented different scientific and clinical backgrounds. MCC members’ diverse knowledge enabled them to hold their assessments of the empirical literature on misophonia to multidisciplinary standards and criteria, as well as relate misophonia to other conditions so that misophonia could be better differentiated from similar disorders.

The primary goal of the Committee was to determine whether or not a consensus definition for misophonia could be developed from the available scientific evidence. The published literature includes various descriptions of misophonia that are based on identifying individuals with misophonia by using different diagnostic questionnaires and measurement tools. While most of these measurement questionnaires and diagnostic checklists have yet to be psychometrically validated, developing diagnostic criteria for misophonia is beyond the scope of the effort undertaken by the Misophonia Consensus Committee.

Finally, the Committee’s assessment of candidate definitional statements is based on the current literature and thus serves as a starting point. As the field’s understanding of misophonia evolves through ongoing research efforts and future scientific inquiry, this body of literature will grow and the definition should be validated, reevaluated, and likely revised.

## Conclusion

The purpose of this project was to determine whether the current body of published literature supported the development of a consensus definition of misophonia. Through the efforts of a Misophonia Consensus Committee using a modified Delphi process, a consensus definition of misophonia was developed from previously published definitional statements that each had at least 80% agreement from Committee members. This definition represents an important first step for researchers and clinicians to progressively build-upon and revise as the body of knowledge in the published scientific literature grows over time. We hope that this consensus definition can bring necessary clarity for individuals experiencing misophonia, the growing community of clinicians who support them, and researchers who seek to better understand this disorder.

## Data Availability

All data generated through this study and used to underscore the study findings can be made available, upon request, by contacting the corresponding author.

## Acknowledgements

The consensus definition project was commissioned by The REAM Foundation through their partnership with the Milken Institute Center for Strategic Philanthropy. The authors thank The REAM Foundation for their strong commitment to the misophonia research community through their development of, and ongoing support for, the Misophonia Research Fund (MRF). The authors also wish to thank the MRF Scientific Advisory Board for prioritizing the development of a foundational understanding of misophonia as one of the earliest MRF strategic initiatives, and for their ongoing support of the consensus definition project.

